# Factors Related to Stress in Children with Online Media Learning Methods during a Pandemic at Jaya Mulya 1 Elementary School, Karawang-Indonesia

**DOI:** 10.1101/2022.04.02.22273339

**Authors:** Ratih Bayuningsih, Fajar Sidik

## Abstract

Stress on students is a condition that occurs when there is pressure at school that makes students feel burdened. This study aims to determine the factors related to stress in students who attend school with online media during the covid-19 pandemic at SDN Jayamulya 1 Kab. Karawang. This study uses a descriptive analytic design with a cross sectional approach with a population of 57 respondents and the sample used is 57 respondents. The results of this study there is a relationship between high achievement pressure factors and stress with a p-value of 0.022 < 0.05, a busy schedule factor with stress with a p-value of 0.012 < 0.05, academic achievement factors with stress with a p-value of 0.000 <0.05, physical demands factor with stress with p-value 0.036 < 0.05.

## INTRODUCTION

On March 24, 2020 the Minister of Education and Culture of the Republic of Indonesia issued Circular Letter Number 4 of 2020 concerning the Implementation of Education Policies in the Emergency Period of the Spread of COVID, in the Circular it was explained that the learning process was carried out at home through online/distance learning starting from kindergarten level up to university level, the online learning process is implemented to provide a meaningful learning experience for students. Studying at home can be focused on life skills education, including regarding the Covid-19 pandemic.

Online learning is the use of the internet network in the learning process. The use of online media or multimedia-based media is one solution to make students able to understand learning materials well. Online learning or distance learning is applied by learning through social media such as WhatsApp, Google Classroom, Google Meet and Zoom.

Research conducted by (Purba, 2020) on the level of stress in students who attend school with online media at Madrasah Aliyah Negeri 2 Model Medan, it was found that 30 students (34.1%) were in the moderate category, most of whom experienced mild stress as many as 22 students (25%), then severe stress and normal stress were 18 students (20.5%). Stress on students can also occur because of the learning system at school. The learning system implemented in schools during the COVID-19 pandemic requires schools to change their learning system with online media regarding the spread of Coronavirus Disease (COVID-19) in Indonesia.

The change from face-to-face learning to online learning that is carried out suddenly makes learning unable to run optimally (Irawati & Jonatan, 2020). The online learning system itself has the advantage that students have the flexibility of learning time, can study anytime and anywhere. Students can interact with teachers with a variety of e-learning applications such as classrooms, video conferences, telephone or live chat, zoom, or via whatsapp groups, but the online learning system also has drawbacks, namely the lack of interaction between students and teachers, or even between students themselves. Students who do not have high learning motivation will tend to fail, where students or teachers are not provided with internet facilities (Amir et al., 2020).

Over time distance learning can have an impact on students’ psychology. Students began to complain about several things, such as network constraints, limited and wasteful data packets because they had to conduct online meetings through the application, the difficulty of working on group assignments, even online learning which had only been done for a few days students had been given many tasks, to the point of stress going to work on tasks that were needed. should be prioritized first. This then has an impact on the mental health of students to cause symptoms of stress. Academic stress on students will arise when expectations for academic achievement increase, assignments that are not in accordance with the student’s capacity, have problems with friends and are bored with learning (Riyandi et al., 2018).

The impact of distance learning is also evident in the students of SDN Jayamulya 1 Kabuaten Karawang, where they feel a level of stress caused by a busy schedule with piled up tasks, lack of rest time, irritability and decreased endurance. Seeing the impact of distance learning that can cause stress levels in children, the authors are interested in researching “Factors related to stress in children with online learning methods at SDN Jayamulya 1, Karawang Regency.”

## METHOD

The design of this research is descriptive analytic with a cross sectional approach. The population in this study were all 5th and 6th graders at SDN Jayamulya 1, Karawang Regency with a total of 57 students. The sampling is done by total sampling. The data collection tool in this study used a questionnaire sheet which was made based on the theory of the variables studied. The questionnaire consists of 2 types, namely: questionnaire A which contains demographic data of respondents consisting of full name, age, and gender, while questionnaire B contains factors related to stress levels in children who go to school with online media during the covid-19 pandemic. consisting of 14 questions.

Data analysis for univariate data uses a frequency distribution to determine the mean, median and mode. Meanwhile, for bivariate data, the data analysis used chi square. The variables in this study were factors related to stress in children using online learning methods at SDN Jayamulya 1, Karawang district. The ethics in this study are: informed consent, anonymity (without a name), and Confidentiality (confidentiality).

## RESULTS

The analysis of the research results was carried out in 2 stages, namely univariate analysis and bivariate analysis. Analysis. The results of the univariate analysis are as follows:

1. Univariate analysis: includes gender, and factors that influence stress levels in children: achievement pressure, busy schedule, academic achievement, physical demands, task demands, role demands and interpersonal demands.
2. Bivariate analysis: to determine the relationship between factors that cause stress in children with online learning methods at SDN Jatimulya 1, Karawang Regency in 2021.

**Table 5.1.**
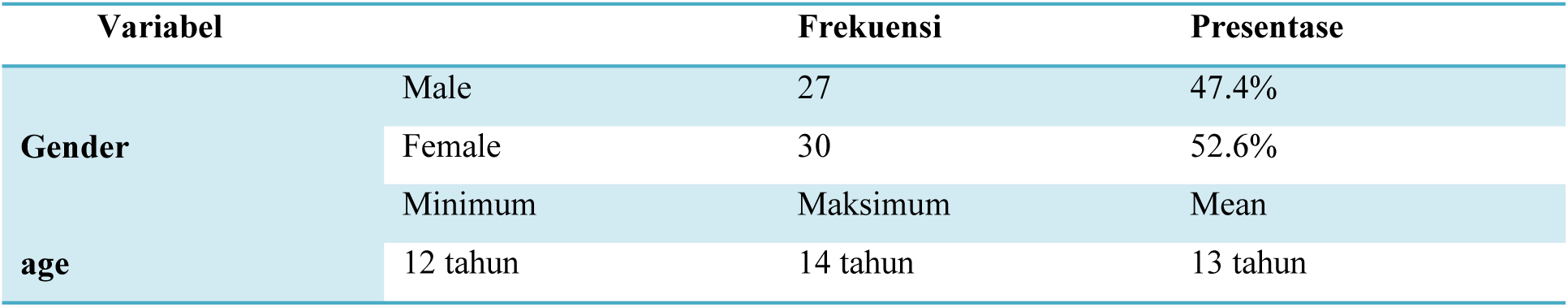
Frequency Distribution by gender and age at SDN Jayamulya 1 Kab. Karawang Tahun 2021

| Variabel |  | Frekuensi | Presentase |
| --- | --- | --- | --- |
| Gender | Male | 27 | 47.4% |
|  | Female | 30 | 52.6% |
| age | Minimum | Maksimum | Mean |
|  | 12 tahun | 14 tahun | 13 tahun |

**Tabel 5.2.** Frequency Distribution by factor related to Stress at children in SDN Jayamulya 1 Kab. Karawang Tahun 2021

| Variable |  | Frekuensi | Prosentase |
| --- | --- | --- | --- |
| Achievement pressure | Exist | 29 | 50.9 |
|  | There is not any | 28 | 49.1 |
| **hectic schedule** | full | 30 | 52.6 |
|  | not full | 27 | 47.4 |
| **Academic achievement** | Not good | 41 | 71.9 |
|  | good | 16 | 28.1 |
| **Physical demands** | bad | 28 | 49.1 |
|  | good | 29 | 50.9 |
| **Task demands** | bad | 26 | 45.6 |
|  | good | 31 | 54.4 |
| **Role Demand** | Tidak baik | 34 | 59.6 |
|  | Baik | 23 | 40.4 |
| **Interpersonal demands** | bad | 29 | 50.9 |
|  | good | 28 | 49.1 |

**Tabel 5.3.**
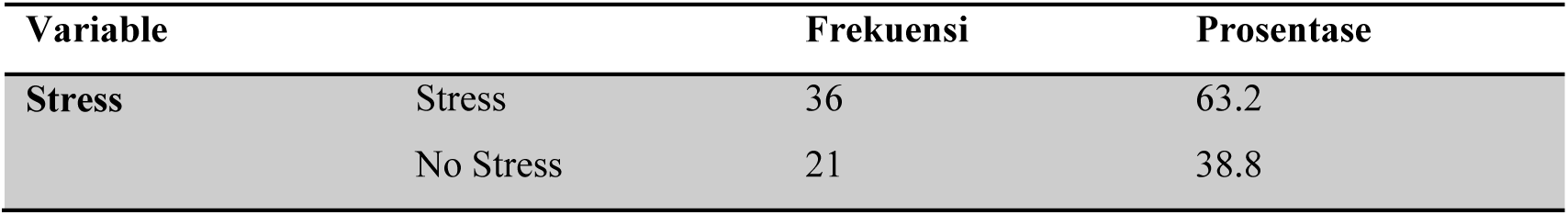
Frequency Distribution by Stress At children in SDN Jatimulya 1 Kab. Karawang Tahun 2021

**Tabel 5.4.** The Relationship of High Achievement Pressure Factors to Stress in Children With Online Media Learning Methods During the Covid-19 Pandemic At SDN Jayamulya 1 Kab. Karawang Year 2021

| Stres |  |  |  |  |  |  | P-Value | OR | 95% CI |
| --- | --- | --- | --- | --- | --- | --- | --- | --- | --- |
| High Performance Pressure | Stres |  | No Stress |  | Total |  |  |  |  |
|  | N | % | N | % | n | % |  |  |  |
| There's Pressure | 23 | 40.4% | 6 | 10.5% | 29 | 50.9% | 0.022 | 4.423 | (1.379-14.190) |
| No pressure | 13 | 22.8% | 15 | 26.3% | 28 | 49.1% |  |  |  |
| Total | 36 | 63.2% | 21 | 36.8% | 57 | 100% |  |  |  |

**Tabel 5.5.** Relationship between busy schedule factors and stress in children with online media learning methods during the Covid-19 pandemic At SDN Jayamulya 1 Kab. Karawang Year 2021

| Stres |
| --- |

| **Hectic Schedule** | Stres |  | No Stres |  | Total |  | P- Value | OR | 95% CI |
| --- | --- | --- | --- | --- | --- | --- | --- | --- | --- |
|  | N | % | N | % | N | % |  |  |  |
| **Hectic Schedule** | 24 | 42.1% | 6 | 10.5% | 30 | 52.6% | 0.012 | 5.000 | (1.547-16.162) |
| **Not Busy Schedule** | 12 | 21.1% | 15 | 26.3% | 27 | 47.4% |  |  |  |
| **Total** | 36 | 63.2% | 21 | 36.8% | 57 | 100% |  |  |  |

**Tabel 5.6.** Relationship between solid academic achievement factors and stress For Children with Online Media Learning Methods During the Covid-19 Pandemic At SDN Jayamulya 1 Kab. Karawang Year 2021

| Stres |  |  |  |  |  |  |  |  |  |
| --- | --- | --- | --- | --- | --- | --- | --- | --- | --- |
| Academic Achievement | Stres |  | No Stres |  | Total |  | P-Value | OR | 95% CI |
|  | N | % | N | % | N | % |  |  |  |
| Bad | 35 | 61.4% | 6 | 10.5% | 41 | 71.9% | 0.000 | 87.500 | (9.679-791.045) |
| Good | 1 | 1.8% | 15 | 26.3% | 16 | 28.1% |  |  |  |
| Total | 36 | 63.2% | 21 | 36.8% | 57 | 100% |  |  |  |

**Tabel 5.7.**
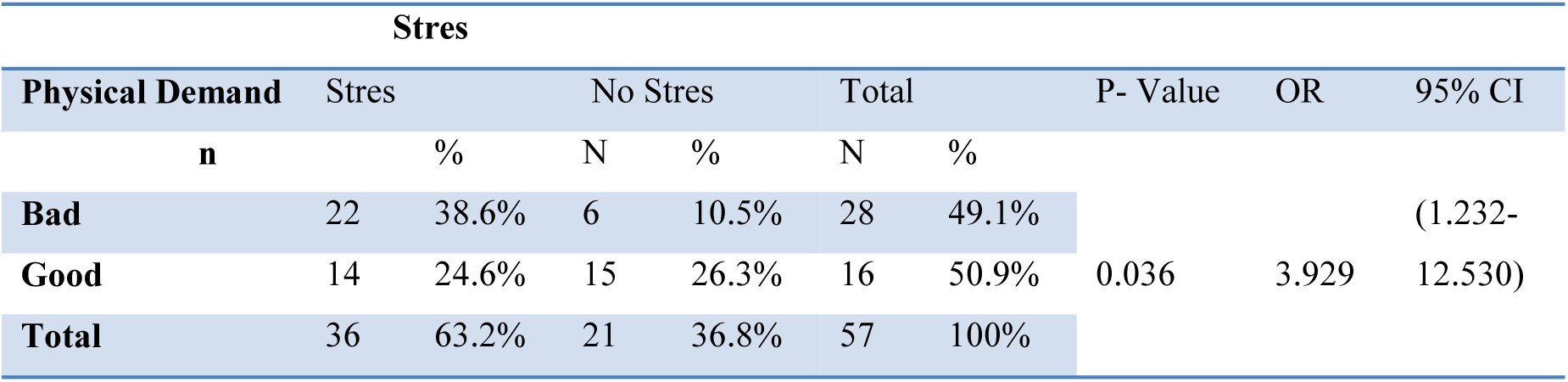
The Relationship of Physical Demand Factors on Stress in Children with Online Media Learning Methods During the Covid-19 Pandemic at SDN Jayamulya 1 Kab. Karawang 2021

**Tabel 5.8.**
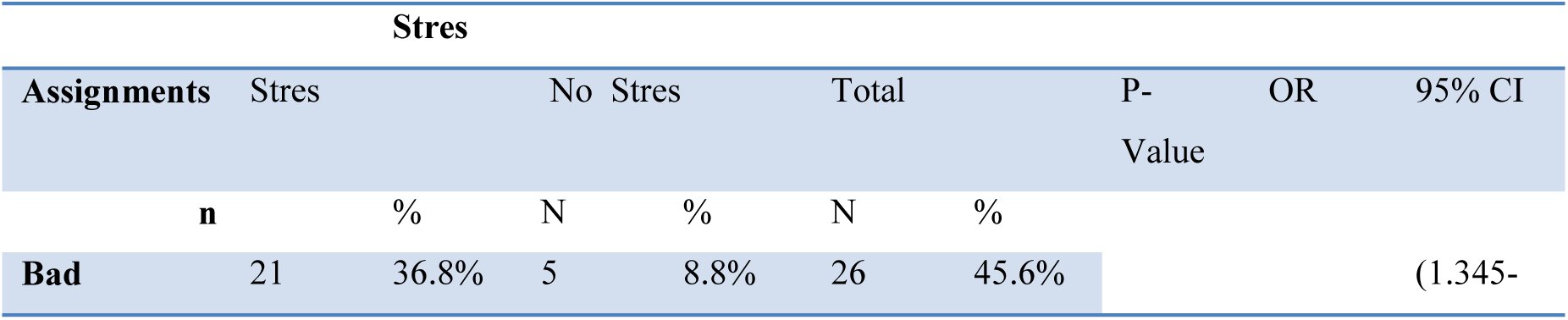
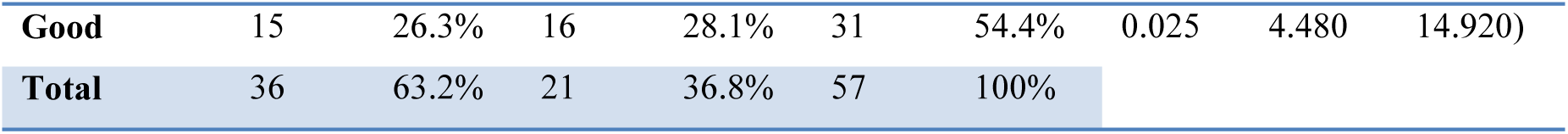
The Relationship of Task Demand Factors on Stress in Children with Online Media Learning Methods During the Covid-19 Pandemic at SDN Jayamulya 1 Kab. Karawang 2021

| Stres |  |  |  |  |  |  | P-Value | OR | 95% CI |
| --- | --- | --- | --- | --- | --- | --- | --- | --- | --- |
| Assignments | Stres |  | No Stres |  | Total |  |  |  |  |
|  | n | % | N | % | N | % |  |  |  |
| Bad | 21 | 36.8% | 5 | 8.8% | 26 | 45.6% |  |  | (1.345- |
| **Good** | 15 | 26.3% | 16 | 28.1% | 31 | 54.4% | 0.025 | 4.480 | 14.920) |
| **Total** | 36 | 63.2% | 21 | 36.8% | 57 | 100% |  |  |  |

**Tabel 5.9.** The Relationship of Role Demand Factors on Stress in Children With Online Media Learning Methods During the Covid-19 Pandemic at SDN Jayamulya 1 Kab. Karawang 2021

| Stres |  |  |  |  |  |  |  |  |  |
| --- | --- | --- | --- | --- | --- | --- | --- | --- | --- |
| Role Demand | Stres |  | No Stres |  | Total |  | P-Value | OR | 95% CI |
|  | N | % | n | % | N | % |  |  |  |
| Bad | 29 | 50.9% | 5 | 8.8% | 34 | 59.6% | 0.000 | 13.257 | (3.614-48.637) |
| Good | 15 | 26.3% | 16 | 28.1% | 31 | 54.4% |  |  |  |
| Total | 36 | 63.2% | 21 | 36.8% | 57 | 100% |  |  |  |

**Tabel 5.10.**
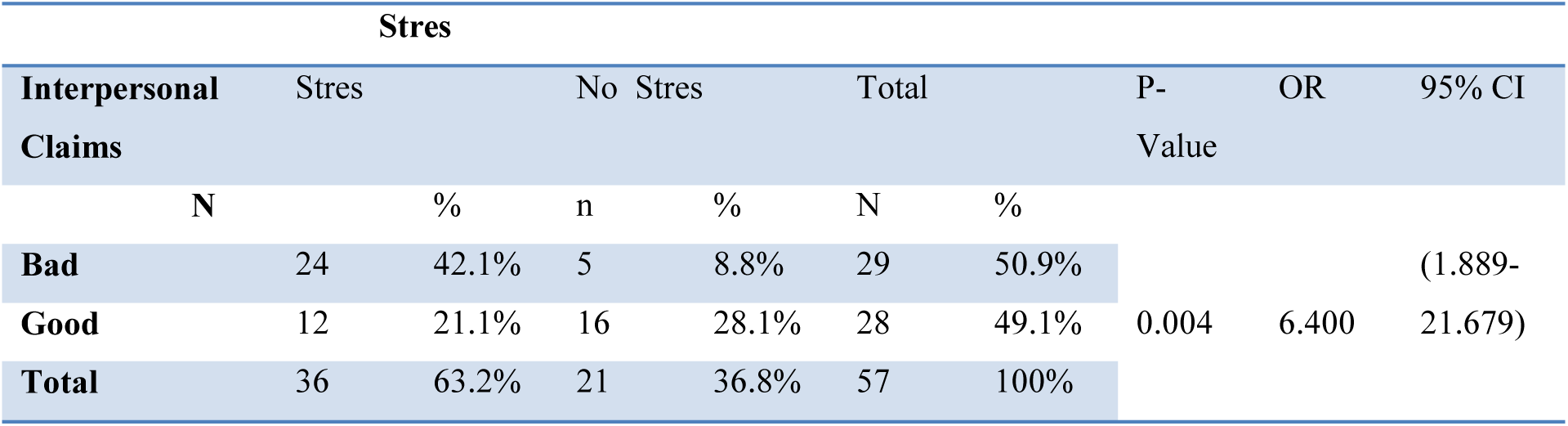
The Relationship of Interpersonal Demand Factors on Stress in Children With Online Media Learning Methods During the Covid-19 Pandemic Period at SDN Jayamulya 1 Kab. Karawang 2021

## DISCUSSION

### The Relationship of High Achievement Pressure Factors to Stress Experienced by Students Using Online Media Methods During the Covid-19 Pandemic Period at SDN Jayamulya. 1 Kab. Karawang

From the statistical test, it was found that Ha was accepted, so it can be concluded that there is a significant relationship between high achievement pressure and stress. From the analysis of risk factors, the OR (Oods Ratio) is 4,423 with 95% Confidence Interval (1,379-14,190). This means that respondents with high achievement pressure have 4,423 times risk of experiencing stress compared to those without high achievement pressure. These results are in line with research conducted by Purba, S (2020) entitled “Stress Levels in Students Who Study with Online Media at Madrasah Aliyah Negeri 2 Medan Model” which states that high achievement pressure is one of the main causes of stress in students.

### Relationship between busy schedule factors and stress experienced by students using online media methods during the Covid-19 pandemic at SDN Jayamulya. 1 Kab. Karawang

From the statistical test, it was found that Ha was accepted, so it can be concluded that there is a significant relationship between a busy schedule and stress. These results are in line with research conducted by Purba, S (2020) entitled “Stress Levels in Students Who Study with Online Media at Madrasah Aliyah Negeri 2 Medan Model” which states that a busy schedule is a source of stress that can occur at school.

### The Relationship between Academic Achievement Factors and Stress Experienced by Students Using Online Media Methods During the Covid-19 Pandemic Period at SDN Jayamulya. 1 Kab. Karawang

Based on the results of the study, it was found that Ha was accepted, it can be concluded that there is a significant relationship between academic achievement and stress. This result is in line with research conducted by Anggrini, D (2021) entitled “Factors triggering stress in high school students during online learning “stated that students would feel stressed because their academic grades fell, it was found that there were 18 students out of 40 students (45%) who felt anxious and afraid when their grades dropped.

### The Relationship between Physical Demand Factors and Stress Experienced by Students Using Online Media Methods During the Covid-19 Pandemic Period at SDN Jayamulya. 1 Kab. Karawang

From the statistical test, it was found that Ha was accepted, so it can be concluded that there is a significant relationship between physical demands and stress. These results are in line with research conducted by (Purba, 2020) entitled “Levels of Stress in Students Who Study with Online Media at Madrasah Aliyah Negeri 2 Medan Model” which states that demands originating from the school’s physical environment include classroom climate conditions, temperature height, lighting and lighting, facilities and infrastructure to support learning and school health.

### The Relationship between Task Demand Factors and the Stress Experienced by Students Using Online Media Methods During the Covid-19 Pandemic Period at SDN Jayamulya. 1 Kab. Karawang

From the statistical test, it was found that Ha was accepted, so it can be concluded that there is a significant relationship between task demands and stress. These results are in line with research conducted by (Anggrini, 2021) entitled “Factors that trigger stress in high school students during online learning” the demands of various learning tasks that cause feelings of stress in students. The indicators are assignments that are done at school and tasks that are done at home, curriculum demands, facing exams or tests, discipline at school, and participating in various extracurricular activities. Students feel anxious if they cannot submit assignments on time.

### Relationship of Role Demand Factors on Stress Experienced by Students Using Online Media Methods During the Covid-19 Pandemic Period at SDN Jayamulya. 1 Kab. Karawang

From the statistical test, it was found that Ha was accepted, so it can be concluded that there is a significant relationship between role demands and stress. The role here is how students can carry out their roles as students well, such as coming to school on time, following teacher suggestions and collecting assignments properly. The results of this study are in line with research conducted by (Anggrini, 2021) entitled “Factors that trigger stress on high school students during online learning” states that students’ anxiety if they can’t submit assignments on time is a factor that causes stress to students, there are 22 students out of 40 students (55%) who feel very anxious when they can’t submit assignments on time.

### Relationship between Interpersonal Demand Factors and Stress Experienced by Students Using Online Media Methods During the Covid-19 Pandemic at SDN Jayamulya. 1 Kab. Karawang

From the statistical test, it was found that Ha was accepted, so it can be concluded that there is a significant relationship between interpersonal demands and stress in elementary school students. This result is in line with the theory put forward by Mathney, 1993 in Palupi, TN (2020) which says that academic stress felt by elementary school students is caused by social stressors, namely stress related to interactions or interpersonal relationships at school, such as interacting with teachers, friends. peers, as well as all forms of student participation in class.

## CONCLUSION

The conclusion of this study is that there are seven factors that influence the stress of students learning to use online media during the COVID-19 pandemic, including:

1. Most of the respondents indicated that the highest gender was female as many as 30 respondents (52.6%).

2. The average age of the respondents shows that the age of the respondents is 13 years.

3. Most of the respondents indicated that the achievement pressure factor as much as 50.9% was a factor causing stress in children

4. Most of the respondents indicated that the busy schedule factor as much as 52.6% was a factor causing stress in children.

5. Most of the respondents indicated that the academic achievement factor as much as 71.9% is a factor causing stress in children.

6. Most of the respondents indicated that the physical demand factor as much as 49.1% can be a cause of stress in children.

7. Most of the respondents indicated that the role demands factor as much as 59.6% can cause stress in children.

8. Some respondents indicated that 45.6% of task demands were the cause of stress in children.

9. Most of the respondents indicated that 50.5% of interpersonal demands were the cause of stress in children.

## Data Availability

All data produced in the present work are contained in the manuscript

## Notes

### Competing Interest Statement

The authors have declared no competing interest.

### Funding Statement

This study did not receive any funding

### Author Declarations

Ethics committee of STIKes Horizon Karawang gave ethical approval for this work

